# Do Rib-Based Anchors Impair Chest Wall Motion in Early Onset Scoliosis (EOS)?

**DOI:** 10.1101/2024.05.01.24306556

**Authors:** Yubing Tong, Jayaram K. Udupa, Joseph M. McDonough, Lipeng Xie, Caiyun Wu, Yusuf Akhtar, Mahdie Hosseini, Mostafa Alnoury, Shiva Shaghaghi, Samantha Gogel, David M. Biko, Oscar H Mayer, Drew A. Torigian, Patrick J. Cahill, Jason B. Anari

**Affiliations:** Medical Image Processing Group, Department of Radiology, University of Pennsylvania, Philadelphia, PA, 19104, United States; The Wyss/Campbell Center for Thoracic Insufficiency Syndrome, Children’s Hospital of Philadelphia, Philadelphia, PA, 19104, United States

**Keywords:** early onset scoliosis, chest wall motion, main thoracic curve, dynamic magnetic resonance imaging

## Abstract

**Purpose:** There is a concern in pediatric surgery practice that rib-based fixation may limit chest wall motion in early onset scoliosis (EOS). The purpose of this study is to address the above concern by assessing the contribution of chest wall excursion to respiration before and after surgery.

**Methods:** Quantitative dynamic magnetic resonance imaging (QdMRI) is performed on EOS patients (before and after surgery) and normal children in this retrospective study. QdMRI is purely an image-based approach and allows free breathing image acquisition. Tidal volume parameters for chest walls (CWtv) and hemi-diaphragms (Dtv) were analyzed on concave and convex sides of the spinal curve. EOS patients (1-14 years) and normal children (5-18 years) were enrolled, with an average interval of two years for dMRI acquisition before and after surgery.

**Results:** CWtv significantly increased after surgery in the global comparison including all EOS patients (p < 0.05). For main thoracic curve (MTC) EOS patients, CWtv significantly improved by 50.24% (concave side) and 35.17% (convex side) after age correction (p < 0.05) after surgery. The average ratio of Dtv to CWtv on the convex side in MTC EOS patients was not significantly different from that in normal children (p=0.78), although the concave side showed the difference to be significant.

**Conclusion:** Chest wall component tidal volumes in EOS patients measured via QdMRI did not decrease after rib-based surgery, suggesting that rib-based fixation does not impair chest wall motion in pediatric patients with EOS.

## Introduction

Early onset scoliosis (EOS) is defined as a spine or chest wall deformity diagnosed prior to the age of 10 [1]. As severe deformity progresses in this patient population, children may go onto develop thoracic insufficiency syndrome (TIS), defined as the inability of the thoracic cavity to support normal respiration or lung growth [2,3]. Treatment for these children involves preservation of pulmonary function through restoration of a less deformed thoracic cavity, promoting thoracic growth, and limiting early spine fusion [4]. Those tasks may be accomplished through a variety of methods; primarily through distraction-based growth friendly instrumentation via traditional growing rods (TGR), vertical expandable prosthetic titanium ribs (VEPTR), or magnetically controlled growing rods (MCGR) [5–9]. Additional techniques such as growth guidance or the Shilla using the spine or the pelvis as anchor points also exist to manage these complex deformities [10,11].

An area of clinical equipoise in the EOS community is the type of proximal anchor used in a posterior distraction-based construct. Surgeons have various fixation options that can be broken up into either spine-based or rib-based anchor points. Biomechanical studies exist reporting on fixation strength for various proximal anchor constructs, but a large majority of the studies evaluate deformity correction percentage and complication rate which comprises mostly infections or proximal anchor failure [12–18].

There is a concern in pediatric spine deformity surgery that rib-based fixation may limit chest wall motion. This philosophy stems from the fusion mass created by device migration, the law of diminishing returns and auto fusion identified in EOS at the time of final fusion, and the complex thoracic cage osteotomies needed at time of graduate final fusion surgery needed to correct severe deformity [19–21].

However, no published literature exists that directly answers the question as to whether rib-based anchors impair chest wall motion in EOS. Unfortunately, until recently there was no practical methodology to evaluate the regional contributions of the various components of the respiratory cycle [22, 23]. In this study, we utilize a recently developed methodology, quantitative dynamic magnetic resonance imaging (QdMRI), to explore the answer to this clinical question. QdMRI [24, 25] is an imaging-based non-invasive methodology that provides quantitative information about regional respiratory function. Images are obtained during tidal breathing at rest, without the need for breath-holding, maximal expiratory effort, or other external monitoring devices, making this technique extremely practical for clinical use, even when patients are unable to follow specific breathing instructions [22, 26]. Other major strengths of QdMRI are that it can be performed in very young EOS patients [22] with high spatial and temporal resolution, affords large field of view that covers the entire thorax and abdomen, and offers sufficient image quality to utilize automatic artificial intelligence (AI)-based image segmentation of the lungs and other anatomical structures of interest [27–29].

We hypothesize that children with EOS managed with posterior distraction-based surgery and rib based proximal anchors will not have impairment of the chest wall contribution to the respiratory cycle post-operatively, and thus not experience a clinically significant chest stiffening.

## Materials & Methods

### Subject cohorts and image data sets

This study received Institutional Review Approval through the Children’s Hospital of Philadelphia (CHOP) and University of Pennsylvania, along with Health Insurance Portability and Accountability Act waiver. The healthy children’s data were acquired through an ongoing prospective research study protocol while all EOS patient data related to rib-based surgery were retrieved from a retrospective study protocol. We excluded the dynamic MRI scans from normal subjects or EOS patients with significant body movement during scanning or with obvious image artifacts. Our primary focus was on chest wall and hemi-diaphragm excursions (on concave and convex sides of the spinal curve separately) during respiration before and after surgical intervention, and we compared their properties in EOS patients to those of age and gender matched controls.

A total of 289 dynamic thoracic MRI scans (49 from EOS patients before surgery, 49 from same EOS patients after VEPTR surgery, and 191 from normal children) were obtained and utilized in this study. For each subject, we utilized the 3D MR images at end expiration (EE) and end inspiration (EI) from the 4D (3D + time) constructed image representing one breathing cycle, leading to a total of 578 3D MR images that were analyzed in this study. **Table 1** lists the age and gender demographics of both EOS patients and normal children. The average time interval between the first dMRI and the dMRI after treatment is ∼2 years. As defined by the Scoliosis Research Society (SRS), four major spinal curves can be defined from frontal radiographs [30]: proximal thoracic curve (PTC), main thoracic curve (MTC), thoracolumbar curve (TLC), and lumbar curve (LC). The concave or convex side is defined according to one of the four major curves and the curve apex orientation. In this study, we utilized the major spinal curve information derived from frontal radiographs but did not utilize kyphosis or lordosis information derived from lateral radiographs.

**Table 1.** Number of EOS patients (pre-& post-operatively) and normal control subjects and their age (mean/standard deviation) considered in this work.

|  |  |  |  |
| --- | --- | --- | --- |
|  | EOS Pre-operative | EOS Post-operative | Normal |
| Female | 22 (3.7/3.5) | 22 (6.5/3.6) | 97 (12.2/3.7) |
| Male | 27 (3.4/3.5) | 27 (5.4/3.6) | 94 (11.7/3.5) |

### Image analysis

dMRI data acquisition [24], 4D-image construction [25], intensity standardization [31], and lung segmentation techniques [27, 28] can be found in our previous publications highlighting the components of this technology. The following volumetric parameters were collected and analyzed separately: right lung tidal volume (RLtv), left lung tidal volume (LLtv), right hemi-diaphragm titdal volume (RDtv), left hemi-diaphragm tidal volume (LDtv), right chest wall tidal volume (RCWtv), and left chest wall tidal volume (LCWtv). Additional assessments included the diaphragm-to-chest wall tidal volume ratio (Dtv/CWtv) on the concave and convex sides of the spinal deformity separately, as well as the percent of volume changes following surgery as r = [(volume _post-surgical_ – volume _pre-surgical_)/ volume _pre-surgical_] × 100%. On average, there was a two-year age interval between dMRI scans of EOS patients before and after surgery. To account for growth-related change and focus on purely surgery-related change, age correction was performed on EOS patient data pre-operatively by first fitting a linear function of tidal volume of the structure of interest (lung, chest wall, hemi-diaphragm) as a function of age using normal children’s measurements, and then using that function to estimate the tidal volume value at the same age of the same EOS patient after surgery.

### Statistical analysis

A two-tailed paired t-test was performed to check for statistically significant differences before and after surgery, and a two-tailed unpaired t-test was used to compare EOS patients with healthy controls. The statistical toolbox of MATLAB (R2019b, The Mathworks Inc., Natick, Massachusetts) was employed for statistical analyses. A p value of <0.05 was considered to denote statistical significance.

## Results

Tidal volume comparisons in EOS patients before and after surgery with and without age correction and without distinguishing between concave and convex sides of the spinal curve are shown in Table 2. Without age correction, all parameter tidal volumes increased after surgery (p<0.05). Both left and right chest wall tidal volumes significantly increased after surgery by 50.66% and 41.18% (without age correction), respectively, and by 17.19% and 24.71% (with age correction), respectively. Left and right Dtv (with age correction) did not significantly differ before and after surgery (p>0.05).

**Table 2.** Global comparisons of tidal volume (in cc) of EOS patients before and after surgery without distinguishing concave and convex sides of spinal curve. Each cell shows mean (upper number) and standard deviation (SD) (lower number) values. P values (pre-surgical vs. post-surgical) are shown using paired t-testing. Ratio r = ((post-pre)/pre) × 100%.

| Table 2. Global comparisons of tidal volume (in cc) of EOS patients before and after surgery without distinguishing concave and convex sides of spinal curve. Each cell shows mean (upper number) and standard deviation (SD) (lower number) values. P values (pre-surgical vs. post-surgical) are shown using paired t-testing. Ratio r = ((post-pre)/pre) x 100%. |  |  |  |  |  |  |  |  |  |  |  |  |
| --- | --- | --- | --- | --- | --- | --- | --- | --- | --- | --- | --- | --- |
|  | Without age correction |  |  |  |  |  | With age correction |  |  |  |  |  |
| EOS | LLtv | RLtv | LCWtv | RCWtv | LDtv | RDtv | LLtv | RLtv | LCWtv | RCWtv | LDtv | RDtv |
| Pre | 34.96<br>22.69 | 53.78<br>38 | 21.68<br>15.75 | 28.63<br>20.83 | 13.38<br>8.27 | 25.23<br>18.80 | 43.27<br>22.36 | 65.15<br>37.83 | 27.86<br>15.82 | 32.41<br>20.87 | 21.47<br>9.3 | 38.19<br>19.16 |
| Post | 51.17<br>27.24 | 79.25<br>45.4 | 32.65<br>17.95 | 40.42<br>21.44 | 20.3<br>12.43 | 42.34<br>26.58 | 51.17<br>27.24 | 79.25<br>45.4 | 32.65<br>17.95 | 40.42<br>21.44 | 20.3<br>12.43 | 42.34<br>26.58 |
| r (%) | 46.37 | 47.36 | 50.60 | 41.18 | 51.72 | 67.82 | 18.26 | 21.64 | 17.19 | 24.71 | -5.45 | 10.87 |
| p | <0.001 | <0.001 | <0.001 | <0.001 | <0.001 | <0.001 | 0.012 | 0.004 | 0.022 | 0.006 | 0.463 | 0.112 |
| EOS – early onset scoliosis; LLtv – left lung tidal volume; RLtv – right lung tidal volume; LCWtv – left chest wall tidal volume; RCWtv – right chest wall tidal volume; LDtv – left hemi-diaphragm tidal volume; RDtv – right hemi-diaphragm tidal volume. |  |  |  |  |  |  |  |  |  |  |  |  |

Table 3 shows the comparison of post-operative volumes with volumes of age-matched controls. All tidal volumes except LDtv (p<0.001) are close to that of normal children and do not differ significantly from those of control subjects (p>0.05).

**Table 3.** Global comparisons of tidal volume (in cc) of EOS patients after surgery with age and gender matched normal children. Each cell shows mean (upper number) and standard deviation (SD) (lower number) values. P values (pre-surgical vs. post-surgical) are shown using paired t-testing.

|  |  |  |  |  |  |  |  |
| --- | --- | --- | --- | --- | --- | --- | --- |
|  | LLtv | RLtv | LCWtv | RCWtv | LDtv | RDtv | age (years) |
| EOS Post | 66.30<br>31.19 | 109.38<br>55.05 | 42.24<br>20.50 | 52.97<br>24.89 | 25.49<br>14.92 | 60.04<br>32.67 | 9.85<br>2.94 |
| normal children | 75.56<br>22.77 | 110.20<br>36.07 | 45.38<br>11.53 | 47.31<br>16.24 | 43.57<br>13.32 | 76.47<br>25.31 | 9.91<br>2.97 |
| p value | 0.10 | 0.95 | 0.51 | 0.41 | <0.001 | 0.06 | 0.10 |
| EOS – early onset scoliosis; LLtv – left lung tidal volume; RLtv – right lung tidal volume; LCWtv – left chest wall tidal volume; RCWtv – right chest wall tidal volume; LDtv – left hemi-diaphragm tidal volume; RDtv – right hemi-diaphragm tidal volume. |  |  |  |  |  |  |  |

Table 4 presents tidal volume comparisons before and after surgery when considering the directionality of the spinal deformity without age correction (Table 4a) and with age correction (Table 4b). In this analysis we did not identify major spinal curve types. CWtv significantly increased after surgery by 46.44% and 30.88% (without age correction) for concave and convex sides, respectively (Table 4a), significantly increased after surgery by 28.57% (with age correction) for the concave side (p < 0.002), and non-significantly increased after surgery by 14.82% (with age correction) for the convex side (p = 0.059) (Table 4b).

**Table 4a:**
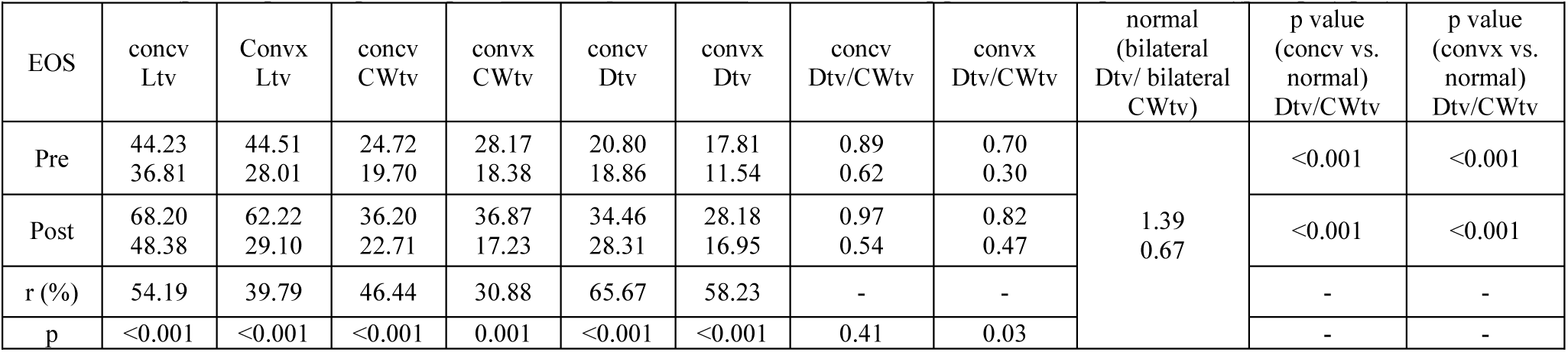
Global comparisons of tidal volumes (in cc) of 49 EOS paired patients before and after surgery distinguishing concave (concv) and convex (convx) sides of spinal curve. Each cell for volumes shows mean (upper number) and standard deviation (SD) (lower number) values. P values (pre-surgical vs. post-surgical without age correction) are shown using paired t-testing. Ratio r = ((post-pre)/pre) × 100%.

**Table 4b.**
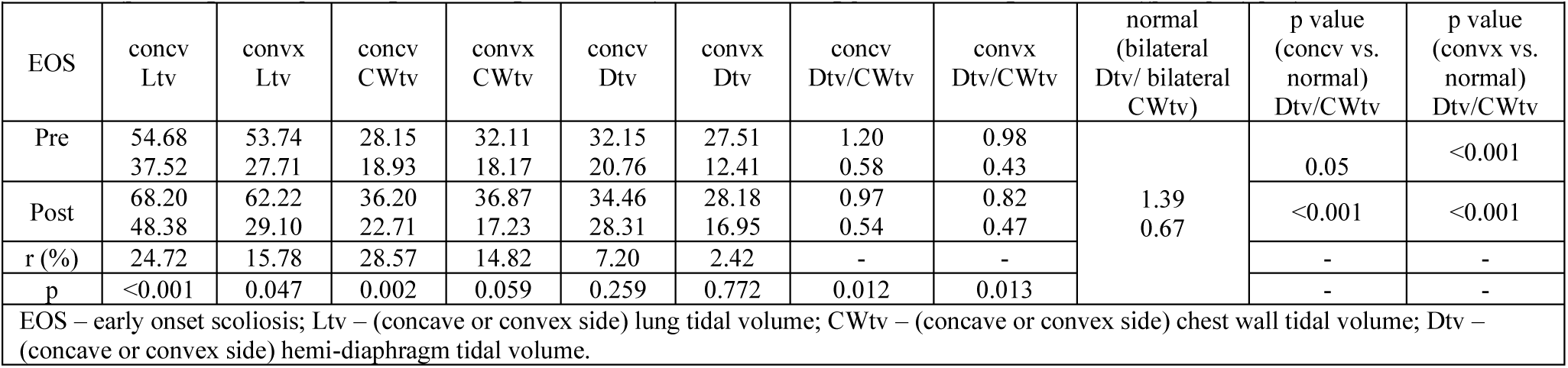
Global comparisons of tidal volumes (in cc) of 49 paired EOS patients before and after surgery distinguishing concave (concv) and convex (convx) sides of spinal curve. Each cell for volumes shows mean (upper number) and standard deviation (SD) (lower number) values. P values (pre-surgical vs. post-surgical with age correction) are shown using paired t-testing. Ratio r = ((post-pre)/pre) × 100%.

Among the 98 studies acquired in 49 paired EOS patients, there were 8 with PTC, 52 with MTC and 20 with TLC and 18 with LC. EOS patients with the MTC type of major spinal curve type formed the largest subgroup with 26 paired EOS patients before and after surgery. Table 5 shows the results of CWtv in this MTC group of EOS patients while considering the concave and convex sides of the spinal curve without age correction (Table 5a) and with age correction (Table 5b). The results of CWtv for the MTC subgroup of EOS patients without distinguishing concave and convex sides of the spinal curve before and after surgery are listed in supplemental Table S1, where CWtv (left or right) significantly increased after surgery (with or without age correction). CWtv significantly increased after surgery for both concave and convex sides without and with age correction (p < 0.05). CWtv increased after surgery by 64.43% (without age correction) and 50.24% (with age correction) on the concave side of the spinal curve, and increased after surgery by 45.35% (without age correction) and 35.17% (with age correction) on the convex side of the spinal curve. Table 4b shows after age correction, the average ratio of tidal volumes Dtv/CWtv in MTC EOS patients after surgery became close to and not significantly different from that of normal children on the convex side of the spinal deformity (1.44±1.10 in MTC EOS patients vs. 1.39±0.67 in normal children (p = 0.78)) compared to that on the concave side of the spinal curve (1.06±0.52 in MTC EOS after surgery vs. 1.39±0.67 in normal children (p = 0.02)). The sub-analysis of CWtv for the 10 patients in the TLC subgroup revealed no significant changes after surgery. TLC and LC subgroups considered together showed no significant changes on CWtv after surgery; see Table S2 for more details.

**Table 5a.**
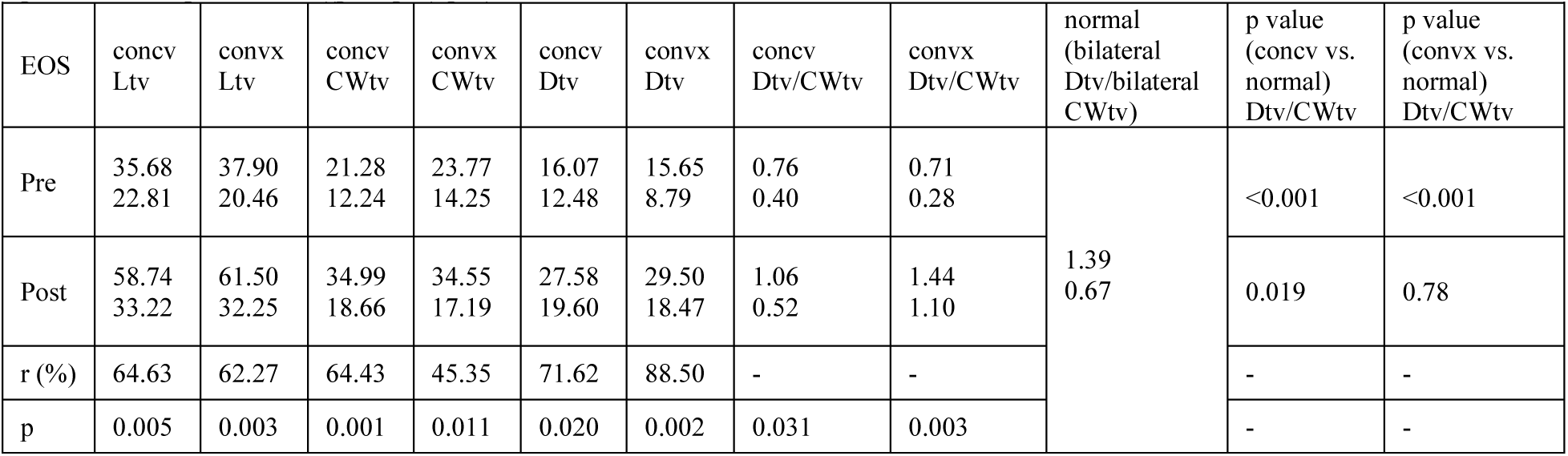
Global comparisons of tidal volumes (in cc) of MTC subset of 26 paired EOS patients before and after surgery distinguishing concave (concv) and convex (convx) sides of spinal curve. Each cell for volumes shows mean (upper number) and standard deviation (SD) (lower number) values. P values (pre-surgical vs. post-surgical without age correction) are shown using paired t-testing. Ratio r = ((post-pre)/pre) × 100%.

**Table 5b.**
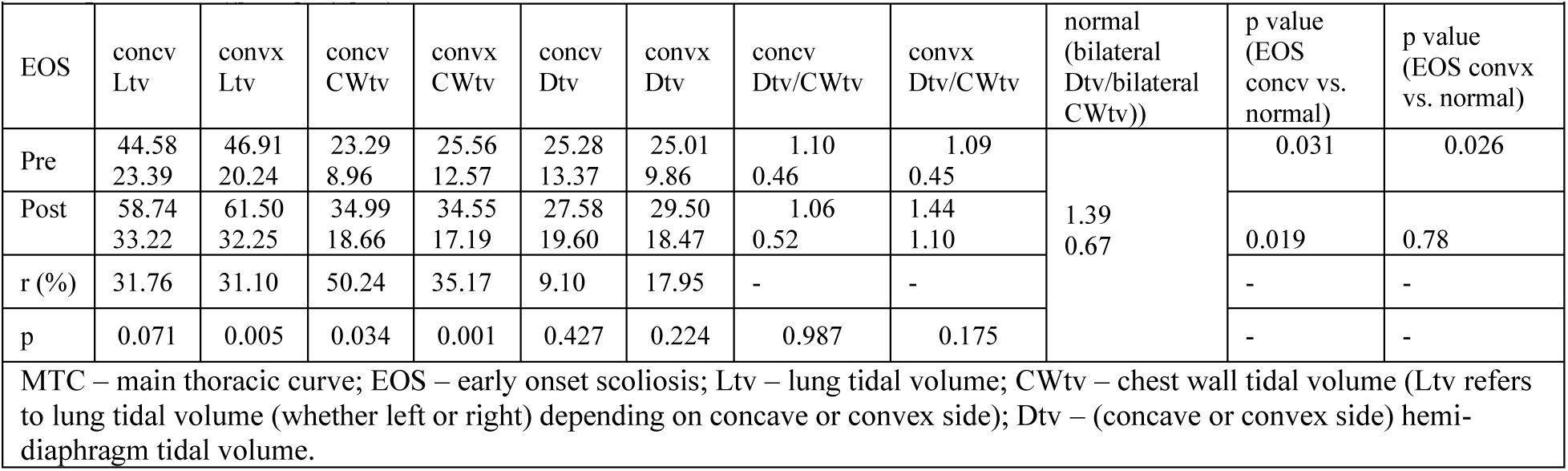
Global comparisons of tidal volumes (in cc) of MTC subset of 26 paired EOS patients before and after surgery distinguishing concave (concv) and convex (convx) sides of spinal curve. Each cell for volumes shows mean (upper number) and standard deviation (SD) (lower number) values. P values (pre-surgical vs. post-surgical with age correction) are shown using paired t-testing. Ratio r = ((post-pre)/pre) × 100%.

Figure 1 depicts a male EOS patient with thoracic spinal dextroscoliosis on the AP radiograph, which also displays 3D MRI surface renditions of his lungs at EE and EI pre-operatively (top row) and post-operatively (bottom row). In this patient, CWtv and Dtv increased after surgery, with bilateral chest wall tidal volumes increasing from 56.71 cc to 127.39 cc (a 124.63% increase), and bilateral hemi-diaphragm tidal volumes combined increased from 91.22 cc to 109.94 cc (a 20.52% increase).

**Figure 1.**
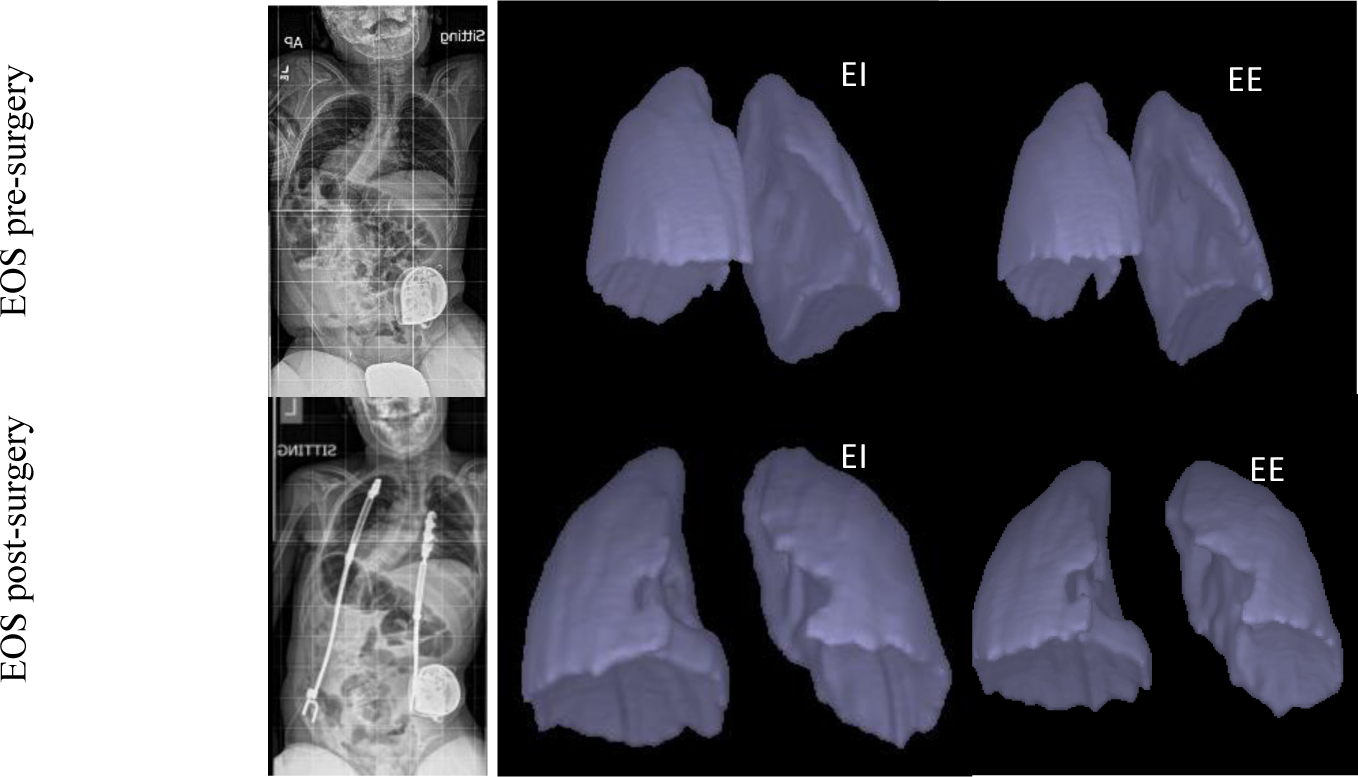
Frontal chest radiographs, and 3D surface rendition results for lungs at EE and EI derived from dMRI for one male EOS patient with leftward concavity of thoracic spine before surgery (the first row) and after surgery (the second row).

## Discussion

Pediatric spine deformity surgeons have a variety of operative techniques to control early onset scoliosis to prevent the development of thoracic insufficiency syndrome. Both Redding and Wang recently concluded that it is important to monitor the impact of different surgical strategies on pulmonary development including the use of various surgical devices and anchor points [32, 33]. However, little data exists, and hence information regarding the long-term changes in lung function during spinal surgical and non-surgical treatments for EOS must be expanded upon [33]. In the current study, we investigated how rib-based fixation affects chest wall motion and thus its contribution to respiratory tidal volume in pediatric patients with EOS by utilizing the recently developed QdMRI methodology [25,26]. We obtained measurements from both the concave and convex sides of the spinal deformity to assess the contribution of chest wall to respiration on its own, relative to the diaphragm, and in comparison, to normal children, before and after surgical intervention.

We observed that globally all tidal volumes of the chest walls and hemi-diaphragms significantly increased after surgery (Table 2). When comparing the post-operative volume contributions from the chest wall to age matched controls we do not see a statistically significant difference (p=0.51 and p=0.41, for left and right chest wall tidal volumes, respectively) (Table 3). This data indicated that the operative intervention utilizing rib-based anchors for EOS in this cohort can return the chest wall contributions of lung volume in the respiratory cycle to near normal, despite the anchor points being attached to the ribs. These results are important to the pediatric spine deformity surgeon as it provides support for proximal anchor attachments to the ribs given the known problematic migration of spine-based pedicle screw anchors in growing constructs [34, 35].

Granular analysis looking at concave and convex sides of the spinal deformity showed that the concave side of the deformity exhibited a greater increase in CWt, Dtv, & the ratio of Dtv/CWtv contributions post-operatively than pre-operatively (Table 4a & 4b) when compared to the convex side. The sub-cohort of MTC, which represents the majority of our cases and is the dominant phenotype by the Lenke classification for adolescent idiopathic scoliosis, exhibited greater increase for CWtv, Dtv, and Dtv/CWtv (Table 5).

Other methods exist for assessing chest wall motion in humans, but no study reached definitive conclusion regarding the effect of posterior-based distraction instrumentation on the respiratory cycle when anchored to the ribs [36–40]. Seddon et al suggested using physical devices placed on the chest wall in conjunction with imaging devices, such as MRI, to measure lung volume, and requiring breathing controls during image acquisition [41]. Such techniques would not be practical for use in EOS patients given their young age and often difficulty with following instructions. Additional studies in the literature compared scoliosis patients via 2D cross-sectional lung displacements or 3D volumes with normal subjects, and concluded that chest wall motion in scoliosis patients is worse than in normal subjects, but this is the first report documenting restoration of normal function post-operatively and regain of function compared to normative control in a patient population that is extraordinarily difficult to assess pulmonary function. Early reports highlighting this novel technology rib-based anchors & the respiratory cycle in children with thoracic insufficiency provides the pediatric spine community an opportunity to answer many pulmonary based questions in a heterogenous patient population that is historically difficult to study. The additional value of this technology lies in the glossary of normative subjects available for comparison, providing surgeons with a baseline to compare to when discussing operative intervention and outcomes with parents.

This study is not without limitation. One limitation of this study is the small number of EOS patients with major spinal curve types beyond MTC such as proximal thoracic, thoracolumbar, or lumbar curves, as not all EOS patients have the same curve type. Another limitation is the small numbers of male and female patients, which precluded gender specific comparisons with curve types, although our normal cohort had roughly equal and large number of male and female subjects. The greatest limitation is the fact that in EOS the most common implant used today is the MCGR, which would preclude a child from getting the QdMRI during treatment, and thus would only allow for post-fusion imaging [42, 43]. Additional limitations include that all data set stems from a single center. Lastly, while the results may demonstrate no detriment to chest wall mobility post-implant, concerns about the possibility of rib-based anchors driving chest wall stiffness cannot be completely dismissed until this cohort is followed to maturity or the time of definitive fusion as stiffness may appear with time. Despite these limitations, in this first study, our results suggest that chest wall motion is not impaired in pediatric patients with early onset scoliosis after rib-based fixation.

## Conclusions

The novel technique of quantitative dynamic magnetic resonance imaging demonstrates that rib-based fixation does not appear to decrease the contribution of chest wall function to the respiratory cycle with chest wall tidal volumes significantly increasing post-operatively for both concave and convex sides of EOS patients with a main thoracic scoliosis and even approaching that of normative age matched controls.

## Data Availability

All data produced in the present study are available upon reasonable request to the authors

## Acknowledgement

This research was supported by a grant R01HL150147 from the National Institutes of Health.

## Supplemental materials

Table S1: Global comparisons of tidal volumes (in cc) of MTC subgroup of EOS patients before and after surgery without distinguishing concave and convex sides of spinal curve. Each cell shows mean (upper number) and standard deviation (SD) (lower number) values. P values (pre-surgical vs. post-surgical) are shown using paired t-testing. Ratio r = ((post-pre)/pre) × 100%.

Table S2: Global comparisons of tidal volumes (in cc) of TLC+LC subgroups of EOS patients before and after surgery without distinguishing concave and convex sides of spinal curve. Each cell shows mean (upper number) and standard deviation (SD) (lower number) values. P values (pre-surgical vs. post-surgical) are shown using paired t-testing. Ratio r = ((post-pre)/pre) × 100%.

