## Supplemental Table S1 for "Do Rib-Based Anchors Impair Chest Wall Motion in Early Onset Scoliosis (EOS)?"

**Table S1**: **Global comparisons of tidal volumes (in cc) of MTC subgroup of EOS patients before and after surgery <u>without distinguishing concave and convex sides of spinal curve</u>. Each cell shows mean (upper number) and standard deviation (SD) (lower number) values. P values (pre-surgical vs. post-surgical) are shown using paired t-testing. Ratio r = ((post-pre)/pre) x 100%.**

| | Without age correction | | | | | | With age correction | | | | | |
|---|---|---|---|---|---|---|---|---|---|---|---|---|
| **EOS** | LLtv | RLtv | LCWtv | RCWtv | LDtv | RDtv | LLtv | RLtv | LCWtv | RCWtv | LDtv | RDtv |
| **Pre** | 31.63<br>19.05 | 41.94<br>22.87 | 20.64<br>13.15 | 24.41<br>13.25 | 12.28<br>7.90 | 19.45<br>12.01 | 46.91<br>20.24 | 44.58<br>23.39 | 25.56<br>12.57 | 23.29<br>8.96 | 25.01<br>9.86 | 25.28<br>13.37 |
| **Post** | 49.43<br>24.76 | 70.80<br>36.02 | 31.10<br>15.42 | 38.45<br>19.45 | 20.17<br>12.80 | 36.91<br>20.43 | 58.74<br>33.22 | 61.50<br>32.25 | 34.99<br>18.66 | 34.55<br>17.19 | 27.58<br>19.60 | 29.50<br>18.47 |
| **r (%)** | 56.28 | 68.81 | 50.68 | 57.52 | 64.25 | 89.77 | 25.22 | 37.95 | 36.89 | 48.35 | 10.28 | 16.69 |
| **p** | 0.004 | <0.001 | 0.007 | 0.001 | 0.006 | <0.001 | 0.071 | 0.005 | 0.034 | 0.001 | 0.427 | 0.224 |

MTC – main thoracic curve; EOS – early onset scoliosis; LLtv – left lung tidal volume; RLtv – right lung tidal volume; LCWtv – left chest wall tidal volume; RCWtv – right chest wall tidal volume; LDtv – left hemi-diaphragm tidal volume; RDtv – right hemi-diaphragm tidal volume.
