## Supplemental Table S2 for "Do Rib-Based Anchors Impair Chest Wall Motion in Early Onset Scoliosis (EOS)?"

**Table S2**: **Global comparisons of tidal volumes (in cc) of TLC+LC subgroups of EOS patients before and after surgery <u>without distinguishing concave and convex sides of spinal curve</u>. Each cell shows mean (upper number) and standard deviation (SD) (lower number) values. P values (pre-surgical vs. post-surgical) are shown using paired t-testing. Ratio r = ((post-pre)/pre) x 100%.**

| | Without age correction | | | | | | With age correction | | | | | |
|---|---|---|---|---|---|---|---|---|---|---|---|---|
| **EOS** | LLtv | RLtv | LCWtv | RCWtv | LDtv | RDtv | LLtv | RLtv | LCWtv | RCWtv | LDtv | RDtv |
| **Pre** | 41.97<br>27.67 | 74.71<br>48.35 | 27.87<br>19.84 | 40.39<br>27.37 | 16.43<br>8.62 | 35.30<br>23.72 | 50.98<br>27.44 | 87.62<br>46.51 | 35.04<br>20.11 | 44.89<br>26.82 | 24.75<br>11.10 | 50.38<br>22.64 |
| **Post** | 52.87<br>34.27 | 91.35<br>59.15 | 33.88<br>23.39 | 41.71<br>27.03 | 20.77<br>13.78 | 51.92<br>33.74 | 52.87<br>34.27 | 91.35<br>59.15 | 33.88<br>23.39 | 41.71<br>27.03 | 20.77<br>13.78 | 51.92<br>33.74 |
| **r (%)** | 25.98 | 22.27 | 21.53 | 3.28 | 26.37 | 47.08 | 3.72 | 4.26 | -3.32 | -7.07 | -16.10 | 3.06 |
| **p** | 0.015 | 0.087 | 0.042 | 0.770 | 0.102 | 0.008 | 0.681 | 0.683 | 0.697 | 0.473 | 0.172 | 0.774 |

MTC – main thoracic curve; EOS – early onset scoliosis; LLtv – left lung tidal volume; RLtv – right lung tidal volume; LCWtv – left chest wall tidal volume; RCWtv – right chest wall tidal volume; LDtv – left hemi-diaphragm tidal volume; RDtv – right hemi-diaphragm tidal volume.
